# Global Rearrangement of Degree Centrality Reflects Cognitive Impairment and Fatigue in Multiple Sclerosis

**DOI:** 10.1101/2024.03.14.24304081

**Authors:** Pavel Hok, Quang Thong Thai, Barbora Rehák Bučková, Martin Domin, Kamila Řasová, Jaroslav Tintěra, Martin Lotze, Matthias Grothe, Jaroslav Hlinka

**Author notes:** Corresponding Author: MUDr. Pavel Hok, Ph.D., Department of Neurology, University Medicine Greifswald Ferdinand-Sauerbruch-Str. 1, 17475 Greifswald, Germany, URL: https://www2.medizin.uni-greifswald.de/neurolog. Both authors contributed equally to this work (Matthias Grothe, Jaroslav Hlinka).

## Abstract

**Background and Objectives:** The aim of this secondary data analysis was to determine whether multiple sclerosis (MS) is associated with changes in global degree rank order disruption index ( *k_D_*), a graph theory-based functional connectivity measure representing shift in overall distribution of nodal degree centrality. Additionally, we tested the relationship between *k_D_* and MS symptoms (cognitive and motor impairment, fatigue, and global disability).

**Methods:** Global *k_D_* was computed in a pre-existing cross-sectional fMRI dataset and compared between patients with MS (PwMS) and healthy controls (HCs). Group differentiation was tested against other known biomarkers in MS (regional degree centrality, structural MRI with volumetry, diffusion-weighted imaging, lesion mapping) using receiver operating characteristic and logistic regression analysis. Associations between *k_D_* and cognitive processing speed (Symbol Digit Modalities Test), fatigue (Fatigue Scale for Motor and Cognitive Functions), gait (Timed Up and Go Test), and disability (Expanded Disability Status Scale [EDSS]) were evaluated using Spearman correlation coefficient and ordinal regression adjusted for structural imaging, age, sex, and disease duration.

**Results:** Analysis included 56 PwMS and 58 HCs (35/27 women, median age 45.1/40.5 years). Global *k_D_* was lower in PwMS (median −0.30, inter-quartile range [IQR] 0.55) than in HCs (median −0.06, IQR 0.54; *p* = 0.009, Mann-Whitney U test). *k_D_* yielded acceptable differentiation between groups (area under curve 0.64), but did not improve such differentiation on top of structural imaging. Both *k_D_* and regional degree in medial prefrontal cortex (MPFC) were correlated with cognitive decline (*k_D_*: Spearman’s *ρ* = 0.32, *p* = 0.019; MPFC: *ρ* = −0.45, *p* = 0.001, *n* = 55), while *k_D_* was also correlated with fatigue (*ρ* = −0.34, *p* = 0.010, *n* = 56), but not with EDSS (*ρ* = −0.06, p = 0.674, *n* = 56) or gait (*ρ* = −0.18, *p* = 0.211, *n* = 52). *k_D_* significantly explained cognitive impairment (*χ*^2^ = 4.49, *p* = 0.034) and fatigue (*χ*^2^ = 7.18, *p* = 0.007).

**Discussion:** Our data provide evidence that *k_D_* is a potential biomarker of cognitive decline and fatigue. Further cross-validations are required to assess its generalizability.

## 1. Introduction

Disentangling the complex interaction between the structural brain damage due to multiple sclerosis (MS) and the resulting clinical presentation is currently one of the most challenging tasks of the MS research as development of new treatment strategies requires reliable biomarkers and predictors of increasing disability and cognitive decline.^1^ Individual lesions in MS do not necessarily translate directly into clinical deficits, since the interplay of functional reserve, adaptation, and maladaptation results in a non-linear relationship between lesion volume and the resulting disability, a fact known as “clinico-radiological paradox”.^2^

Several MRI modalities have been utilized to account for this mismatch and to model functional deterioration in MS, including advanced lesion imaging techniques, T_1_-weighted structural imaging for volume measurement, diffusion weighted imaging (DWI) for assessment of white matter integrity, and functional MRI (fMRI).^1,3,4^ Especially changes in fMRI-based resting-state functional connectivity (rsFC) may reflect clinical deficits that cannot be explained by structural damage alone.^5^ Due to the high-dimensional nature of raw rsFC data, dimensionality reduction procedures are necessary to obtain practicable biomarkers.^6^ Analysis of rsFC using graph theory framework provides such network-wide numerical indices^7^ suitable for routine applications. Despite known abnormalities in graph-theoretical parameters such as mean degree or eigenvector centrality in MS and their association with global disability,^8,9^ no single reliable and widely used graph-theory-based rsFC biomarker has been incorporated into clinical practice, yet.^1,10^

Besides motor and sensory deficits, MS is frequently associated with cognitive impairment (CI).^11^ Being underrepresented in Expanded Disability Status Scale (EDSS)^12^ and often neglected in routine neurological examination,^11,13^ CI and its imaging biomarkers are attracting increasing attention.^1,11^ In our group, the relationship between CI and cerebellar atrophy,^14^ brain activation during a cognitive task,^15^ or damage to the parietal white matter^16^ has been demonstrated. Recent resting-state fMRI studies utilizing graph-theoretical approaches have identified relationship between CI and decreased overall mean degree^17^ or increased centrality in the default mode network (DMN), accompanied by decreased centrality outside the DMN.^18–23^ Moreover, changes in centrality also seem to precede the actual cognitive decline, indicating its potential utility as a prognostic biomarker.^19^ However, the ultimate goal, i.e., providing individual predictions applicable in day-to-day practice remains far from achieved, urging thus further refinement of imaging and analytical methods.^1^

Whereas a voxel-wise centrality assessment would require considerable time and personal resources, a recently introduced centrality-derived global scalar metric, the degree rank order disruption index (*k_D_*),^24^ could serve as a biomarker reflecting simultaneous focal increases and decreases in degree centrality throughout the brain, requiring less demanding interpretation. The *k_D_* has previously been demonstrated to be associated with brain-wide degree centrality changes in impaired consciousness^25^ and chronic pain,^24^ but so far, it was evaluated neither as a biomarker of global disability nor predictor of CI in MS. The advantage of *k_D_* analysis is the potential ability to process data automatically without selecting a specific region of interest, which may be particularly useful in MS characterized by diffuse brain involvement and highly variable lesion distribution patterns.

Hence, the aim of this study was to assess the potential clinical utility of *k_D_* against established structural and functional imaging biomarkers. We therefore investigated the correlation between the clinical presentation of MS (cognitive performance, global disability and motor performance) and *k_D_* in comparison to multimodal MRI parameters (resting-state fMRI with regional degree assessment, diffusion-weighted imaging [DWI], volumetry, and lesion mapping) in patients with MS and matched healthy controls. The hypotheses were as following: (1) *k_D_*differs between patients with MS (PwMS) and matched healthy controls (HCs); (2a) *k_D_* is superior to the regional degree centrality in the pre-selected regions of interest (ROIs) in differentiating between PwMS and HCs and (2b) improves such differentiation when used in conjunction with established global imaging diagnostic biomarkers of MS (lesion load, global atrophy, global white matter integrity); (3) *k_D_* correlates with deficits in cognitive processing speed in PwMS; (4) *k_D_* provides superior correlation with cognitive processing speed in comparison to (4a) the regional degree centrality in pre-selected ROIs and (4b) improves regression model of cognitive processing speed when added on top of established global imaging diagnostic biomarkers of MS (lesion load, global atrophy, global white matter integrity).

In addition, following exploratory hypotheses were tested to assess associations between *k_D_*and potential confounding factors: (5) *k_D_* correlates with global disability, fatigue, and motor performance (gait) and (6) improves regression models for these clinical outcomes when used jointly with the established global imaging diagnostic biomarkers of MS (lesion load, global atrophy, global white matter integrity); (7) *k_D_* correlates with the global imaging biomarkers (lesion load, global atrophy, global white matter integrity); (8) global imaging diagnostic biomarkers of MS (lesion load, global atrophy, global white matter integrity) differ between PwMS and matched HCs.

## 2. Methods

### 2.1 Study Design and Participant Selection

The secondary analysis was performed on cross-sectional imaging and behavioral data of 65 PwMS and 65 HC matched for age and sex, with participants recruited from multiple sclerosis centers across Czechia. The same cohort has been partially analyzed and published using other methods.^6,26^ Original inclusion criteria were: positive diagnosis of multiple sclerosis;^27^ spastic paraparesis as a prominent clinical feature; stable clinical status for at least three months preceding the study (determined by neurologist); physical ability to undergo clinical testing – consistent with the EDSS score ≤ 7.5. Subjects were excluded in case of missing imaging data or imaging artifacts and conditionally excluded if exceeding the motion outlier criteria (see Section 2.5 Data pre-processing and analysis below). To address potential attrition bias, a sensitivity analysis was performed in a sample including motion outlier subjects.

### 2.2 Clinical Assessment and Questionnaires

Clinical parameters (regressands) comprised an objective assessment of cognitive performance using a Symbol Digit Modalities Test (SDMT)^28^ and possible confounding factors such as fatigue (Fatigue Scale for Motor and Cognitive Functions, FSMC),^29^ global disability (Expanded Disability Status Scales, EDSS),^12^ and motor performance (Timed Up and Go Test, TUG).^30^

### 2.3 Standard Protocol Approvals, Registrations, and Patient Consents

The secondary data analysis was pre-registered at osf.io (https://osf.io/v8ejw). The original study was approved by ethics committee of the Ethics Committee of the Faculty Hospital Královské Vinohrady, approval No. EK-VP/22/0/2014. All patients gave their written informed consent to participate in the study.

### 2.4 MRI Data Acquisition

Imaging was performed using a 3T magnetic resonance scanner (Siemens Trio Tim, Erlangen, Germany) equipped with a 12-channel phased-array head coil. The MRI protocol included blood oxygenation level-dependent (BOLD) resting-state fMRI (repetition time [TR] = 2,500 ms, echo time [TE] = 30 ms, flip angle = 70°, voxel size = 3×3×3 mm, 44 contiguous slices, 240 volumes, acquisition time = 10 min), as well as high-resolution 1-mm T1-weighted and T2-weighted imaging, and 2-mm diffusion-weighted imaging (DWI). Detailed acquisition parameters have been published elsewhere.^6,26^

### 2.5 Data Pre-processing and Analysis

The initial pre-processing consisted of motion correction, correction of susceptibility-induced distortions and normalization to standard space using Advanced Normalization Tools (ANTs, v2.3.5.dev212-g44225).^31^ Brain coverage was evaluated using Mask_explorer.^32^ Next, motion outlier detection, anatomical component-based denoising procedure, and band-pass filtering within the frequency range 0.008 Hz – 0.09 Hz were applied in CONN toolbox v. 21a.^33^ Participants with maximum volume-to-volume displacement exceeding 2 mm and/or mean volume-to-volume displacement exceeding 2 standard deviations above the sample mean (i.e., >0.26 mm) were marked as outliers (*n* = 13). In the main analysis, all remaining steps were performed after excluding the outliers. However, additional sensitivity analysis consisting of the same steps was conducted in a parallel pipeline in the sample with outliers. Please also refer to Supplementary Methods for further details on pre-processing.

Following the pre-processing, subject-specific functional connectivity matrices containing Fisher-transformed Pearson’s *r* coefficients were computed in CONN for 4,632 large voxels created using 6-mm resampling of a common gray matter mask, see Supplementary Methods for more details. Degree centrality was then computed for each subject using a brain connectivity toolbox (BCT, https://sites.google.com/a/brain-connectivity-toolbox.net/bct/) with 10% link density.^24^ Regional degree centrality was extracted by averaging nodal degree from the DMN (4 ROIs),^18,19,21^ basal ganglia (6 ROIs),^19,21^ thalamus, hippocampus, and cerebellum (5 ROIs),^21^ and from the multimodal ROIs explicitly participating in SDMT: superior parietal lobule (2 ROIs), dorsolateral prefrontal cortex (2 ROIs), and anterior cingulate cortex (ACC; 1 ROI),^15^ see Supplementary Fig. S1 and Supplementary Table 1 for detailed ROI description. Finally, *k_D_* was calculated using custom Matlab scripts implementing a modified approach according to Mansour et al.^24^ In brief, *k_D_* was obtained for each individual using a linear regression (y = *k_D_* *x + b), where y = individual nodal degree – mean control group nodal degree, x = mean control group nodal degree, and b = intercept of the regression, see Supplementary Methods for more details.

The pre-processing of T1-weighted and DWI data, calculation of gray matter volume (GMV) as a measure of cortical atrophy and extraction of fractional anisotropy (FA) as a measure of white matter integrity, as well as ROI definition for GMV and FA is described elsewhere.^6^ Finally, the lesion load (LL) was calculated using the Lesion Segmentation Tool (LST, www.statistical-modelling.de/lst.html with lesion prediction algorithm.^34^

### 2.6 Statistical Analysis

Initially, normality was assessed for all continuous variables using Kolmogorov-Smirnov test. Non-parametric tests were applied in case normality was violated. Additionally, regressors considerably deviating from normal distribution (LL) were log-transformed prior to any subsequent analysis to meet regression model assumptions. Demographic variables were compared between groups using Fisher’s exact test and Mann-Whitney U test. Pairwise deletion was applied in case of missing clinical data. A summary of all variables and outcome measures is provided in Supplementary Table 2.

The hypotheses were evaluated using following tests: (1) Group differences in *k_D_*(primary outcome measure) between PwMS and HCs were assessed using a Mann-Whitney U test. (2) Differentiation between PwMS and HC based on *k_D_* was assessed using (2a) area under the receiver operating characteristic (ROC) curve (AUC) and compared with differentiation based on regional degree centrality in each ROI using pair-wise asymptotic significance. For comparison with structural imaging parameters (2b), multiple logistic regression *χ*^2^ statistics was used to test whether model with *k_D_*, GMV, FA, and log(LL) significantly improved differentiation between PwMS and HCs compared to a nested model without *k_D_* as a regressor. The correlations between (3) *k_D_* and SDMT, (4a) regional degree centrality and SDMT, as well as auxiliary correlations between *k_D_*and (5) additional clinical variables (EDSS, FSMC, TUG) or (7) structural imaging parameters (GMV, FA, LL) were evaluated using Spearman’s rank correlation coefficient. For correlations of regional degree centrality (4a), Bonferroni-Holm correction for multiple comparisons across 18 ROIs was applied (α = 0.0028). To test whether *k_D_* improves regression of clinical scores on top of structural imaging, ordinal regression models including *k_D_*, GMV, FA, log(LL), age, sex, and disease duration as regressors of (4b) SDMT, (6a) EDSS, (6b) FSMC, and (6c) TUG were compared to nested models without *k_D_* by means of likelihood ratio test. (8) Finally, differences in all imaging parameters (regional degree centrality, global GMV, global FA, and lesion load) were assessed using multiple Mann-Whitney U tests with Bonferroni-Holm correction for multiple comparisons. All tests were performed using SPSS v29.0.1.1 (IBM, Armonk, NY, USA). The p < 0.05 was considered significant. One-tailed tests were used where superiority was assumed by the hypotheses or statistics with one-tailed distribution was employed (2a, 2b, 4b, and 6), with two-tailed tests applied otherwise. Details on figure preparation are provided in Supplementary Methods.

### 2.7 Post-hoc Analyses

In order to visualize local contributions to significant global correlations between *k_D_* and clinical scores, a post-hoc voxel-wise analysis was performed in randomise, part of FSL v. 6.0.3.^35^ First, nodal degree centrality was back-projected to the original 6-mm voxels. Next, general linear model with two-sample t-test (group differences) and regression contrast was employed to evaluate the correlation using non-parametric threshold-free cluster enhancement (TFCE) correction for multiple comparisons with 10,000 permutations and family-wise error corrected alpha = 0.05.

In case of significant correlations with EDSS, FSMC or TUG, correlation with regional degree in the 18 pre-defined ROIs was additionally assessed using Spearman rank correlation to allow further interpretation.

### 2.8 Power Analysis

No *k_D_*data in PwMS for a power analysis were available. Based on a published difference in *k_D_*between patients and HC of |*k_D_*| = 0.21 in a different patient cohort,^24^ analysis results in a minimum sample of 12 participants to achieve power to detect a significant correlation of 90%. With the existing data set (*n* = 64) differences of down to |*k_D_*| = 0.09 can be identified with the same power of 90%.

### 2.9 Data Availability

Data not provided in the article because of space limitations may be shared (anonymized) at the request of any qualified investigator for purposes of replicating procedures and results, upon signing a data sharing agreement. Part of the dataset (imaging data for 60 PwMS with the respective global disability and motor scales) is publicly available in an on-line repository at https://osf.io/p2kj7/. The custom Matlab script for *k_D_* calculation is available at https://github.com/pavelhok/calculate_kd/tree/MS-project.

## 3. Results

### 3.1 Study Sample

Out of the original sample of 65 PwMS and 65 HC, one PwMS and one HC were excluded due to missing data (incomplete field of view and susceptibility artifact), another PwMS was excluded due to a suspected vascular lesion. In the remaining sample, 13 subjects with excessive motion levels were identified (7 PwMS and 6 HCs), see Supplementary Fig. S2 for inclusion/exclusion diagram. Here, only results in 56 PwMS and 58 HCs after excluding motion outliers are reported (“final” sample), whereas results in the sample with outliers are provided in Supplementary Results. While both analyses yielded mostly similar results, two differences are explicitly stated below. Demographic details of the “final” sample and summary statistics for clinical parameters are provided in Table 1.

**Table 1.**
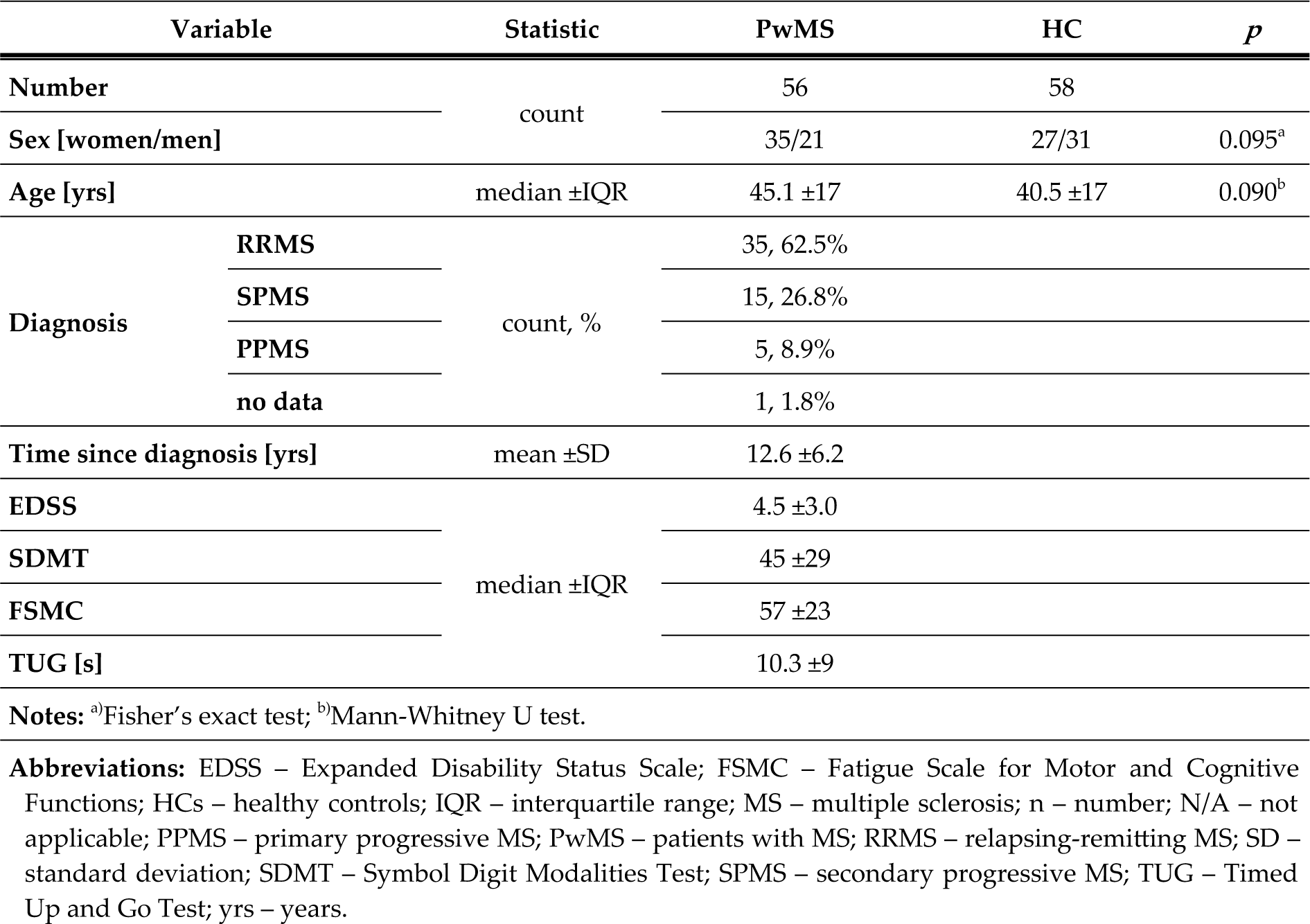
Demographic and clinical data.

### 3.2 Group Differences and Differentiation between PwMS and HCs

PwMS showed significantly lower *k_D_* compared to HCs (PwMS: median = −0.298, inter-quartile range [IQR] = 0.549; HCs: median = −0.058, IQR = 0.542; *p* = 0.009, Mann-Whitney U test), see Fig. 1. Underlying raw degree centrality data are summarized in Supplementary Fig. S3.

**Fig. 1.**
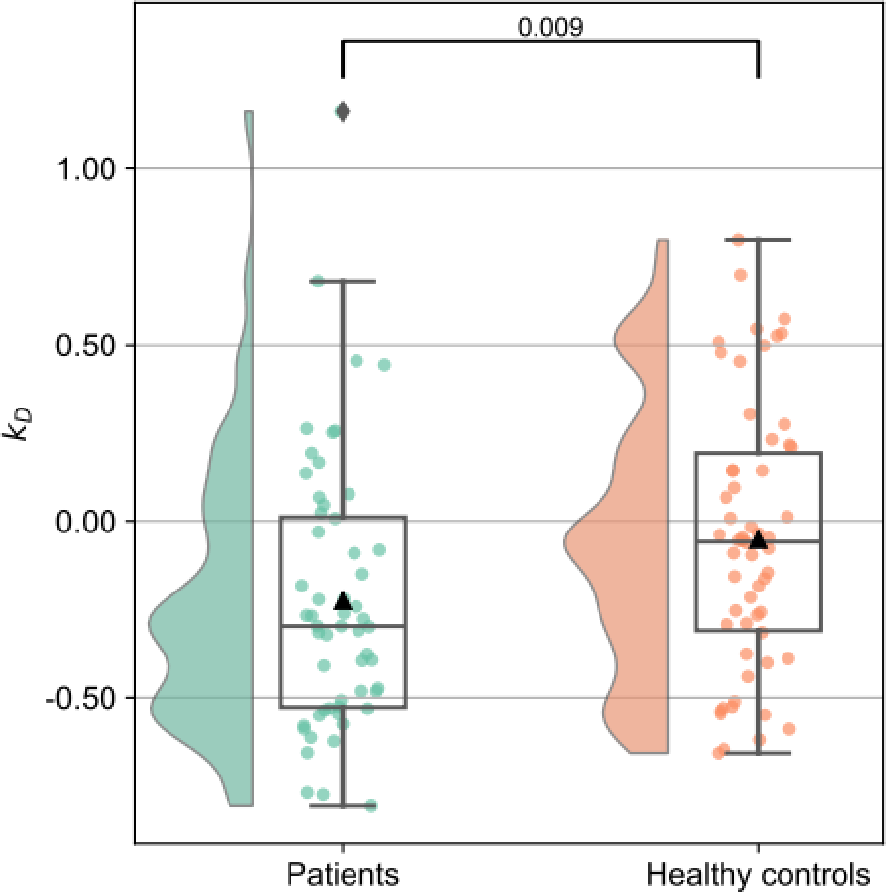
Group differences in degree rank order disruption index (*k_D_*). Raincloud plots illustrating individual *k_D_* values and distribution. Patients with multiple sclerosis are shown in green, healthy controls in red. Mann-Whitney U test *p* value is provided in annotation.

The ROC analysis for differentiation between PwMS and HCs yielded significant above-chance area under curve (AUC) for *k_D_* (AUC = 0.642, *p* = 0.007, two-tailed asymptotic significance for null hypothesis AUC = 0.5), the left lateral parietal portion of the DMN (DMN-LLP; AUC = 0.671, *p* = 0.001), and the ACC (AUC = 0.619; *p* = 0.026), see Fig. 2 and Supplementary Table 3. In pair-wise comparisons, AUC for *k_D_* was significantly higher than AUC for 6 ROIs and did not significantly differ from the remaining ROIs (Supplementary Table 3).

**Fig. 2.**
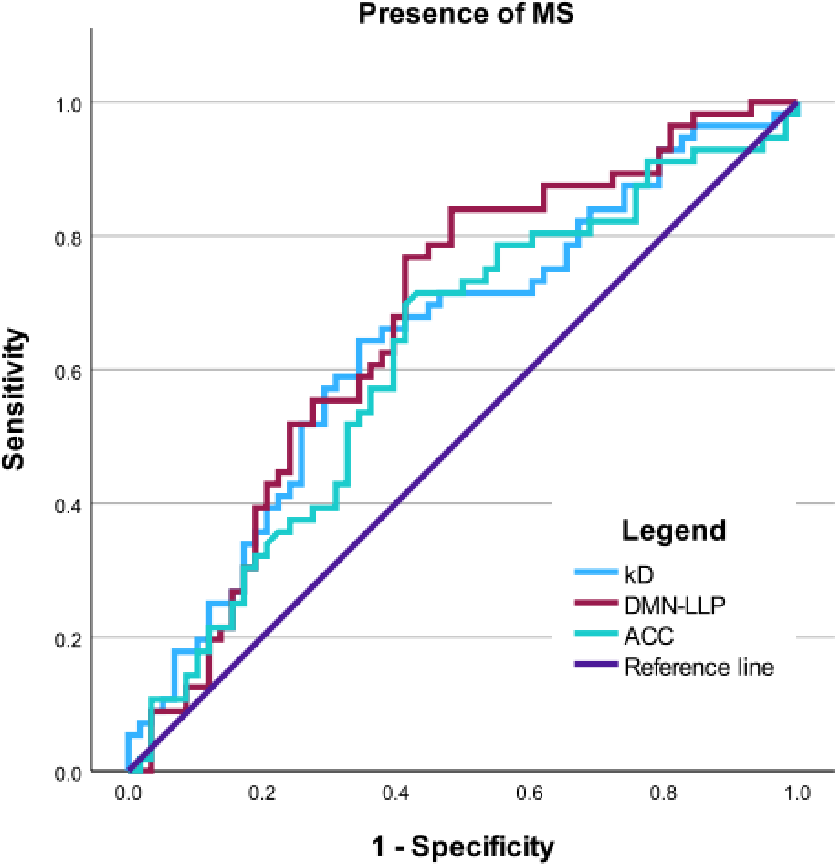
Receiver operating characteristic (ROC) analysis. ROC curves for differentiation between patients with multiple sclerosis (MS) and healthy controls using degree rank order disruption index (*k_D_*; area under curve [AUC] = 0.642; *p* = 0.007, two-tailed asymptotic significance for null hypothesis AUC = 0.5, uncorrected); the left lateral parietal portion of the default mode network (DMN-LLP; AUC = 0.671; *p* = 0.001), and the anterior cingulate cortex (ACC; AUC = 0.619; *p* = 0.026).

We observed no significant improvement in a multiple logistic regression model differentiating between PwMS and HCs after adding *k_D_* as an additional regressor on top of GMV, FA, log(LL) (*χ*^2^ step = 0.007, *p* = 0.934).

### 3.3 Correlation with Cognitive Processing Speed

We detected a significant correlation between *k_D_* and SDMT (Spearman’s *ρ* = 0.32, p = 0.019, *n* = 55, Fig. 3). For the regional degree centrality, significant correlation with SDMT was observed in the medial prefrontal part of the DMN, yielding slightly higher effect size than the *k_D_*, see Table 2. An ordinal regression model including GMV, FA, log(LL), age, sex, and years since diagnosis as regressors of SDMT score was significantly improved after adding *k_D_* (*χ*^2^ step = 4.49, *p* = 0.034, likelihood ratio test, see Table 3). In contrast, neither significant correlation with SDMT nor improvement of regression model for SDMT were observed in analysis with motion outliers (see Supplementary Results).

**Fig. 3.**
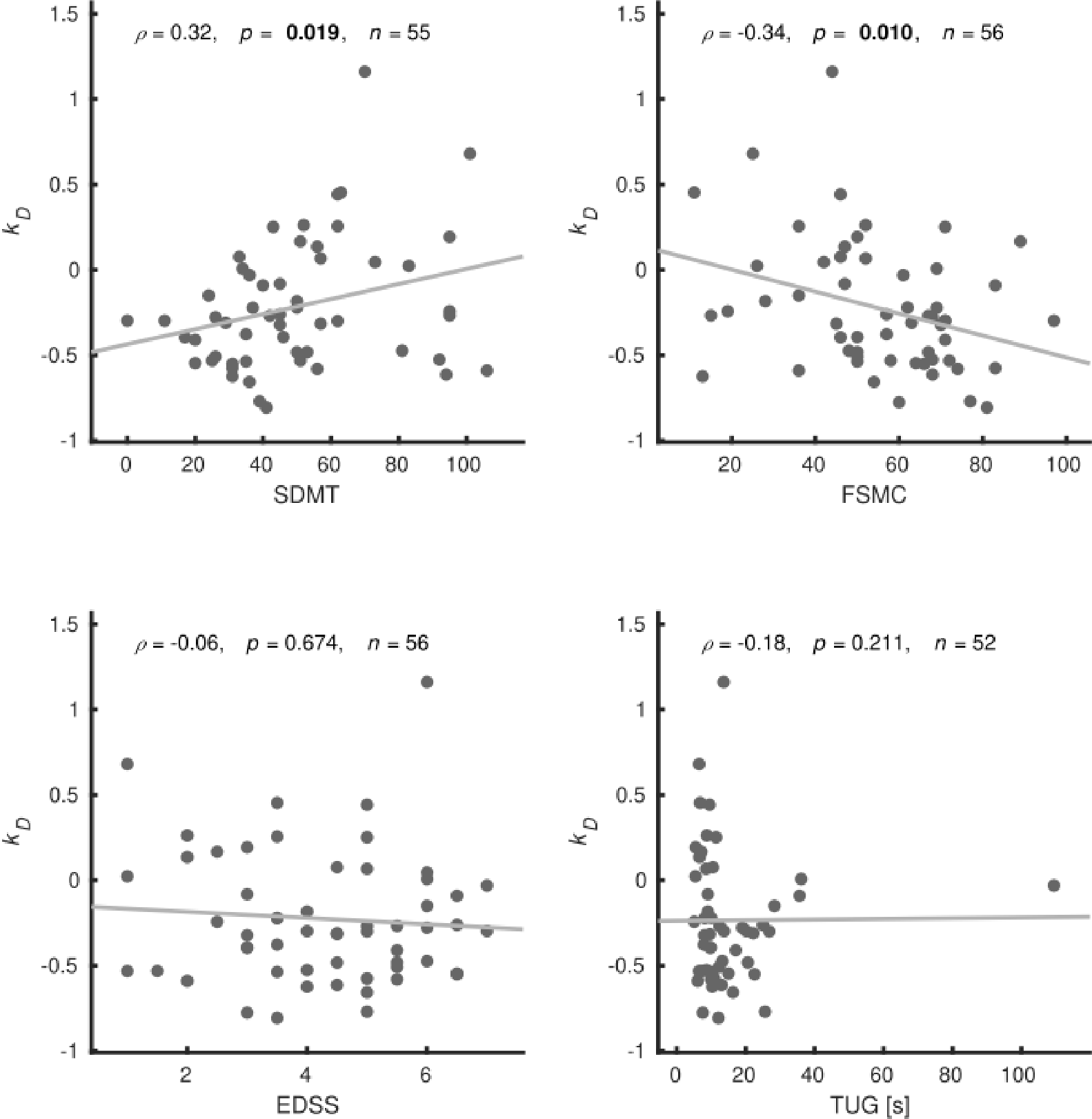
Correlation between *k_D_* and clinical scores. Scatter plots illustrating relationship between the degree rank order disruption index (*k_D_*) and cognitive processing speed (Symbol Digit Modalities Test, SDMT), global disability (Expanded Disability Status Scales, EDSS), fatigue (Fatigue Scale for Motor and Cognitive Functions, FSMC), and motor performance (Timed Up and Go Test, TUG). Spearman’s rank correlation coefficient ( *ρ*), two-tailed uncorrected significance, and number of valid observations are provided.

**Table 2.**
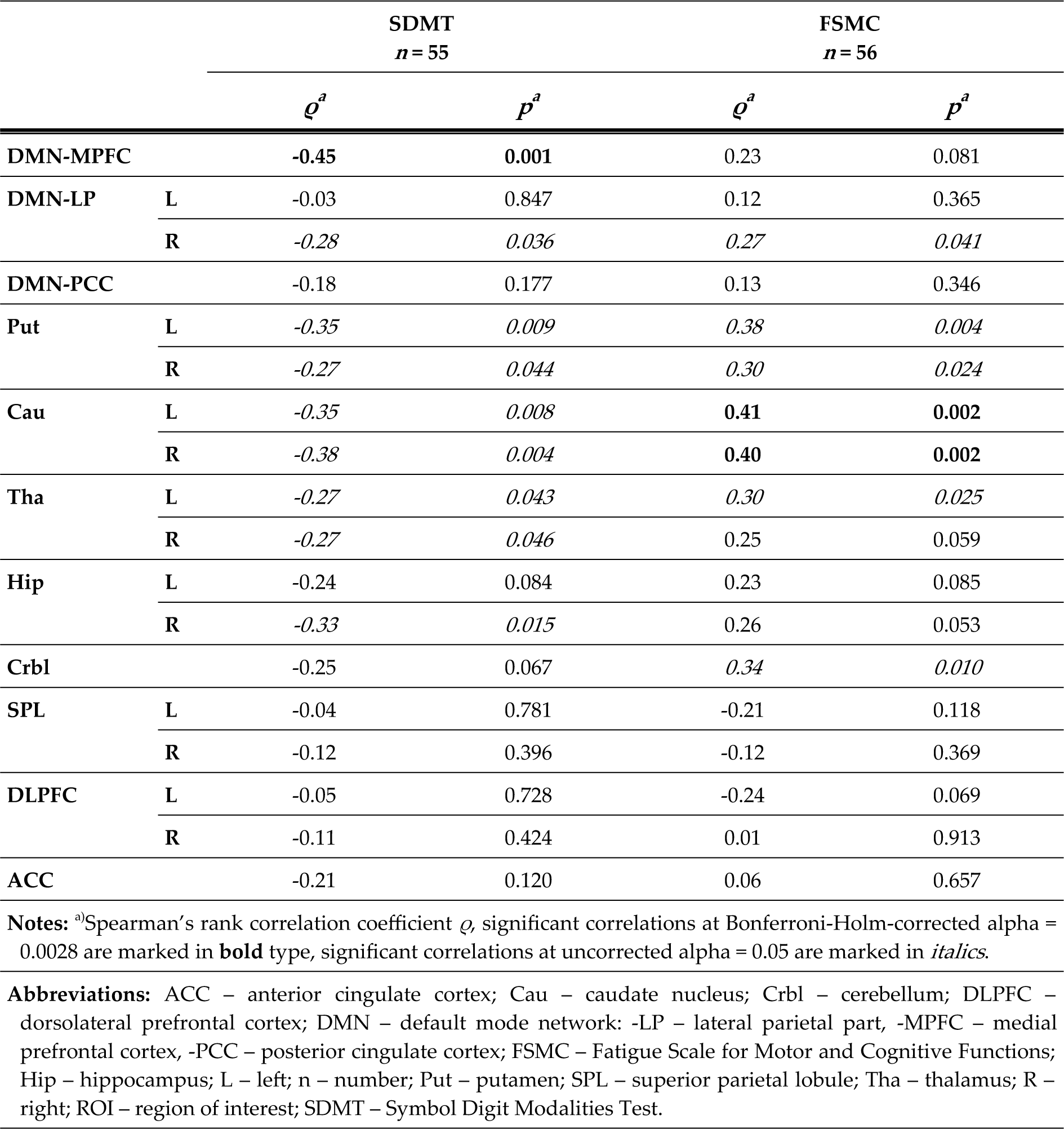
Correlation between regional degree and clinical scores.

|  |  | SDMT<br>*n* = 55 |  | FSMC<br>*n* = 56 |  |
| --- | --- | --- | --- | --- | --- |
| | | $\rho^a$ | $p^a$ | $\rho^a$ | $p^a$ |
| DMN-MPFC |  | -0.45 | 0.001 | 0.23 | 0.081 |
| DMN-LP | L | -0.03 | 0.847 | 0.12 | 0.365 |
|  | R | -0.28 | 0.036 | 0.27 | 0.041 |
| DMN-PCC |  | -0.18 | 0.177 | 0.13 | 0.346 |
| Put | L | -0.35 | 0.009 | 0.38 | 0.004 |
|  | R | -0.27 | 0.044 | 0.30 | 0.024 |
| Cau | L | -0.35 | 0.008 | 0.41 | 0.002 |
|  | R | -0.38 | 0.004 | 0.40 | 0.002 |
| Tha | L | -0.27 | 0.043 | 0.30 | 0.025 |
|  | R | -0.27 | 0.046 | 0.25 | 0.059 |
| Hip | L | -0.24 | 0.084 | 0.23 | 0.085 |
|  | R | -0.33 | 0.015 | 0.26 | 0.053 |
| Crbl |  | -0.25 | 0.067 | 0.34 | 0.010 |
| SPL | L | -0.04 | 0.781 | -0.21 | 0.118 |
|  | R | -0.12 | 0.396 | -0.12 | 0.369 |
| DLPFC | L | -0.05 | 0.728 | -0.24 | 0.069 |
|  | R | -0.11 | 0.424 | 0.01 | 0.913 |
| ACC |  | -0.21 | 0.120 | 0.06 | 0.657 |

Notes: <sup>a</sup>Spearman's rank correlation coefficient $\rho$ , significant correlations at Bonferroni-Holm-corrected alpha = 0.0028 are marked in **bold** type, significant correlations at uncorrected alpha = 0.05 are marked in *italics*.
**Abbreviations:** ACC – anterior cingulate cortex; Cau – caudate nucleus; Crbl – cerebellum; DLPFC – dorsolateral prefrontal cortex; DMN – default mode network; -LP – lateral parietal part, -MPFC – medial prefrontal cortex, -PCC – posterior cingulate cortex; FSMC – Fatigue Scale for Motor and Cognitive Functions; Hip – hippocampus; L – left; n – number; Put – putamen; SPL – superior parietal lobule; Tha – thalamus; R – right; ROI – region of interest; SDMT – Symbol Digit Modalities Test.

**Table 3.**
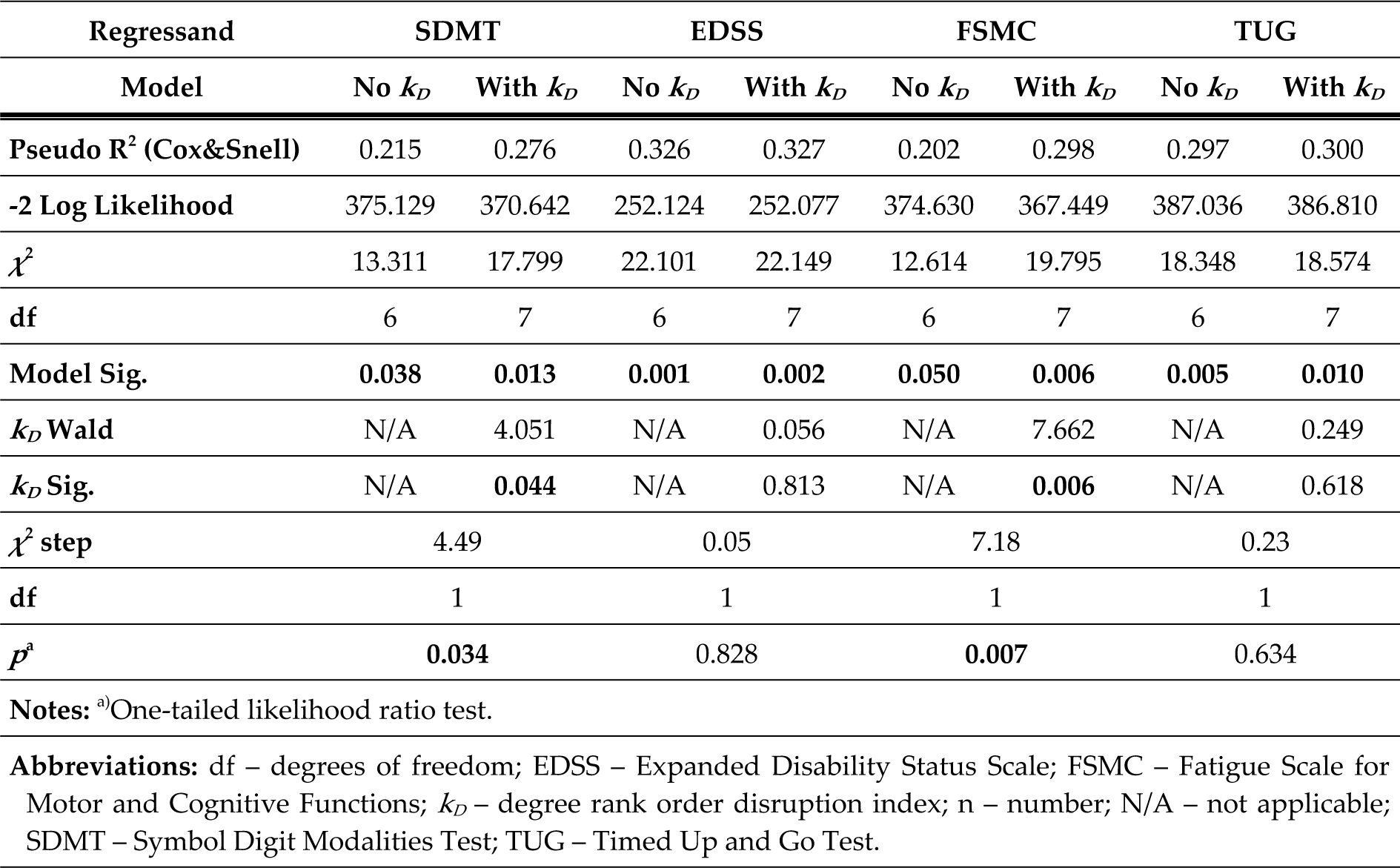
Ordinal regression of clinical scores.

### 3.4 Correlation with Fatigue, Global Disability, and Motor Performance

We detected a significant correlation between *k_D_* and FSMC (Spearman’s *ρ* = −0.34, *p* = 0.010, *n* = 56), but not for EDSS (Spearman’s *ρ* = −0.06, *p* = 0.674, *n* = 56) or TUG (Spearman’s *ρ* = −0.18, *p* = 0.211, *n* = 52), see Fig 4. In ordinal regression, *k_D_* significantly improved the model fit for fatigue (FSMC) when added on top of GMV, FA, log(LL), age, sex, and years since diagnosis, but not for EDSS or TUG (Table 3).

**Fig. 4.**
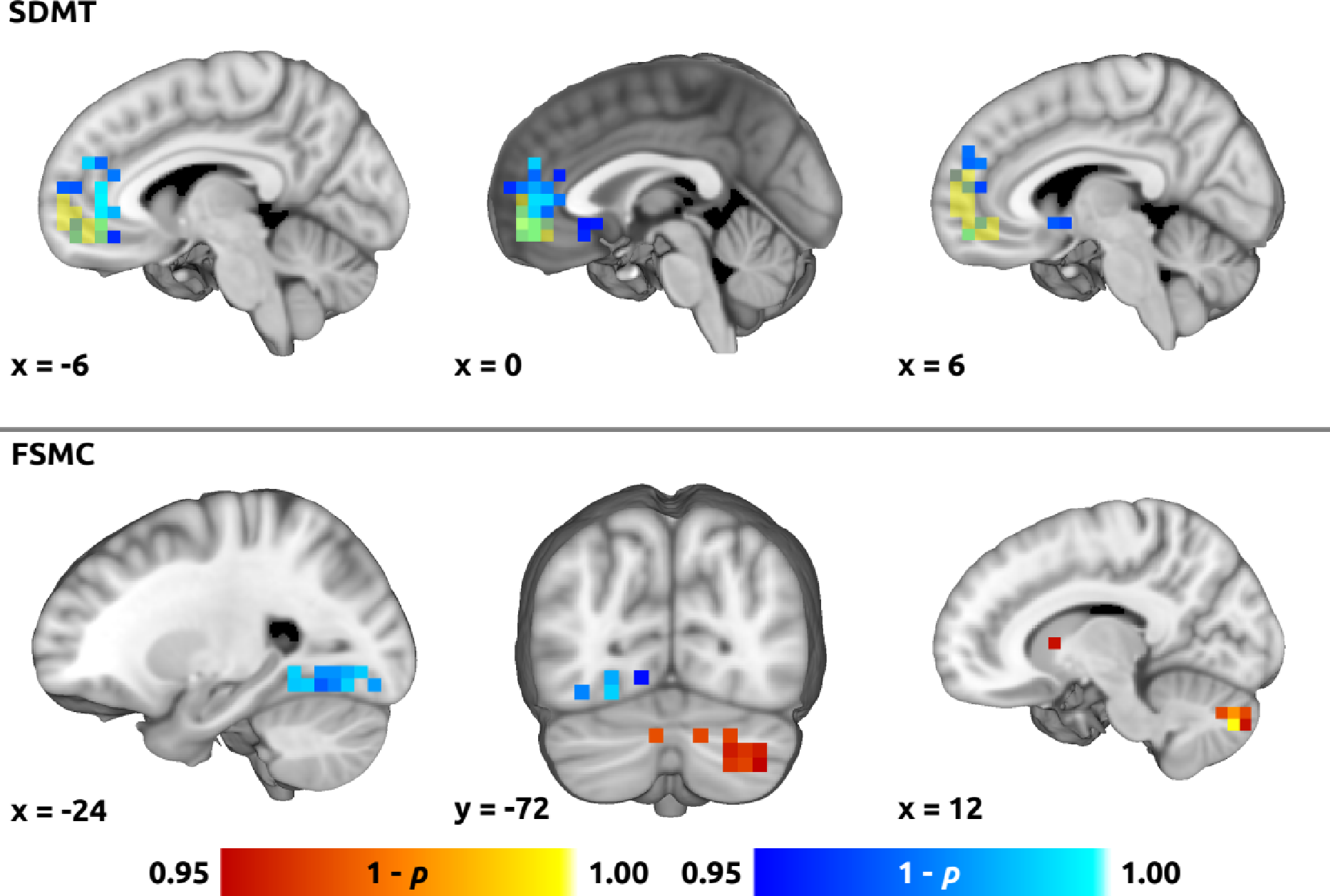
Voxel-wise correlation between degree and clinical scores. Color overlays illustrating spatial distribution of correlation of the voxel-wise degree centrality with (**top**) cognitive processing speed (Symbol Digit Modalities Test, SDMT) and (**bottom**) fatigue (Fatigue Scale for Motor and Cognitive Functions, FSMC). Red-yellow and blue-lightblue overlays indicate positive and negative correlation, respectively. Thresholded using non-parametric threshold-free cluster enhancement (10,000 permutations, family-wise error-corrected *p* = 0.05). In the **top** panel, yellow overlay shows overlap with the medial prefrontal cortex default mode network (MPFC-DMN) region of interest.

### 3.5 Relationship between *k_D_* and Structural Imaging Biomarkers

We observed no significant correlation between *k_D_* and structural imaging parameters, i.e., GMV, LL, and global FA, see Supplementary Fig. S4. In analysis with motion outliers, however, *k_D_* was significantly correlated with LL (see Supplementary Results). All structural imaging parameters significantly differed between PwMS and HCs, see Supplementary Table 4.

### 3.6 Post-hoc Analyses

Voxel-wise group differences are illustrated in supplementary Fig. S3. Voxel-wise regression analysis for degree centrality as response variable and SDMT as explanatory variable yielded significant cluster with negative effect mainly in MPFC and ACC, and to a lesser degree in the subcallosal cortex and the right nucleus accumbens, overlapping in part with the DMN-MPFC ROI (Fig. 4 and Fig. S3 for unthresholded data). The regression for FSMC yielded a positive effect in the predominantly right cerebellum, right caudate nucleus, right inferior frontal gyrus and a negative effect in the left temporo-occipital fusiform and lateral occipital cortex (Fig. 4 and Fig. S3 for unthresholded data).

The ROI analysis for FSMC yielded significant correlation in the left and right caudate nuclei (left: Spearman’s *ρ* = 0.41, *p* = 0.002; right: Spearman’s *ρ* = 0.40, *p* = 0.002; *n* = 56, Bonferroni-Holm-corrected across the 18 ROIs, i.e., alpha = 0.0028). See Table 2 for complete results.

## 4. Discussion

The present study aimed to investigate the potential utility of *k_D_* as a new functional imaging biomarker of CI in MS in comparison to regional degree centrality. While *k_D_*in PwMS significantly differed from HCs and was a significant explanatory variable for cognitive processing speed (SDMT) in PwMS, it yielded weaker correlation than the mean degree centrality in the frontal hub (MPFC) of the DMN. In an exploratory analysis, *k_D_* turned out to be a significant explanatory variable for self-reported fatigue. This was not the case for global disability (EDSS) or motor performance (TUG). A post-hoc analysis indicated, that the correlation with fatigue might be driven by degree centrality changes in the cerebellum, basal ganglia (caudate nuclei), and left fusiform gyrus.

### 4.1 Differentiation between PwMS and HCs

Our primary observation of decreased *k_D_* in PwMS captures the global character of changes in nodal centrality (both degree and eigenvector) that have recently been reported on the local and network-wide level in MS.^9,18,20,21,36^ Lower *k_D_* in PwMS suggests less centralized and more diffusely distributed rsFC, which is analogous to a previously described disruption of the rich-club topology of the brain network in MS.^37^ However, our multiple regression analysis indicated that *k_D_*did not improve differentiation between PwMS and HCs when added on top of structural imaging parameters. Taken together with the considerable overlap between *k_D_*distributions in PwMS and HC (Fig. 1), it can be inferred that degree reordering is predominantly driven by factors other than presence of MS. In fact, changes in centrality of the network hubs have been consistently observed to depend on the cognitive status of PwMS.^19–22^ As such, we conclude that changes in *k_D_* are more likely related to the resulting neurological deficits than to the underlying MS-related pathology *per se*.

### 4.2 Correlation with Cognitive Processing Speed

Our next main analysis demonstrated that *k_D_* was a significant explanatory variable for cognitive processing speed when compared to global structural imaging parameters. While the negative correlation between *k_D_* and SDMT is a novel finding, it is in line with previous evidence for weak positive association between SDMT score and rsFC of the peripheral nodes outside the rich club.^37^ The correlation between *k_D_* and SDMT yielded an effect size similar to some previously reported structural imaging biomarkers of cognitive processing speed, including fractional anisotropy in the superior longitudinal fascicle^16^ or gray matter atrophy-based brain age gap,^38^ but was lower than overall effect size in a recent meta-analysis of multimodal structural MRI data.^39^

In our dataset, a superior correlation with SDMT was achieved using regional degree centrality in the MPFC hub of the DMN, yielding an effect size comparable with structural imaging biomarkers.^39,40^ Correspondingly, cognitive impairment in MS has been shown to be associated with increased degree or eigenvector centrality in the DMN^19–23^ and decreased centrality in the visual^20,22,23^ and sensorimotor network,^20^ while eigenvector and degree centrality show high agreement even within the same group.^21^ Nevertheless, our results add novel evidence for the strength of the relationship between degree centrality in the DMN and cognitive processing speed since no such correlation in MS has been reported before.

### 4.3 Role of the DMN in Pathophysiology of Cognitive Deterioration in MS

The ROI and *post-hoc* voxel-wise analyses indicated that the MPFC and ACC (i.e., anterior DMN) provided the highest correlation between degree and cognitive processing speed. The role of DMN in CI remains controversial.^1^ Although rsFC abnormalities in DMN have been considered to be non-specific, i.e., linked to the dysfunction of the entire brain network,^1^ an excessively central and less dynamic DMN has also been proposed to directly hinder externally oriented cognitive processing by superfluous introspective thoughts.^20,41^ In contrast, cognitive rehabilitation has been associated with increased activation and rsFC of the DMN, suggesting its possible participation in compensatory mechanisms.^42^ Additionally, some rsFC studies using the Paced Auditory Serial Addition Test (PASAT), i.e., another frequently employed cognitive test in MS research, also point to dysfunction of DMN and subcallosal cortex.^36,40^ Hence, graph-theoretical measures extracted from anterior portion of the DMN are potential future candidates for even more accurate biomarkers of cognitive processing speed than *k_D_*, although studies using more elaborate cognitive testing are warranted to assess their specificity with respect to other cognitive domains.

### 4.4 Association with Fatigue

Our exploratory analyses indicated, that apart from cognitive processing speed, *k_D_*was a significant explanatory variable for the self-reported global fatigue score (FSMC). While cognitive performance and fatigue have been shown to be associated with similar rsFC dysfunctions, such as increased rsFC in posterior DMN and reduced rsFC in the anterior DMN,^43^ our *post-hoc* analyses on ROI and voxel-wise level suggested potential differentiation between CI and global fatigue: Whereas SDMT correlated with average degree in DMN-MPFC, fatigue scores were more strongly associated with degree in the caudate nuclei, cerebellum, and fusiform cortex (Table 2 and Fig. 4).

Although cytokine and neuroendocrine abnormalities have been discussed as potential causes of fatigue in MS,^44,45^ it has also been associated with damage to cortico-subcortical pathways and with particular involvement of prefrontal cortex.^45^ On the computational (metacognitive) level, it has been proposed to result from mismatch between predicted and measured output from cognitive and sensorimotor networks.^1,44^ Our results fit in by emphasizing the role of basal ganglia^46^ and cerebellum, which is involved in maintaining internal forward models and error monitoring.^47^ Future dedicated studies should evaluate the specificity and stability of the here identified biomarkers and their accuracy with respect to the motor and cognitive sub-domains of FSMC.

### 4.5 Relationship with Global Disability and Motor Performance

Despite some evidence for increased eigenvector centrality in the basal ganglia,^19^ DMN,^18,19^ and lower degree in executive control and visual networks^36^ in patients with higher EDSS, we found no association between *k_D_* and global disability (EDSS) and gait (TUG). In contrast, previous machine learning study in the same dataset achieved a medium correlation (not exceeding r = 0.5) with EDSS or TUG using rsFC, white matter integrity and gray matter atrophy features either separately or combined.^6^ We therefore speculate that motor deficits are better explained by local changes in rsFC than by overall network disarray. This is supported by fact that PwMS in our paraparetic cohort had significantly lower degree in supplementary motor area and paracentral lobule than HCs as shown by our *post-hoc* analysis (Supplementary Fig. S4). This may be, at least in part, due to strong influence of strategic lesions in pyramidal or cerebellar pathways (including the spinal cord) on motor performance including gait,^48,49^ which dominates the EDSS score.^12^

### 4.6 Correlation with Structural Imaging Parameters

Our results showed that *k_D_* was largely independent on global structural imaging parameters, such as brain atrophy, global white matter integrity or lesion load, supporting its applicability as an add-on biomarker for some of the clinical scores. In previous studies, however, degree centrality in the thalamus and cerebellum correlated with the lesion load,^18^ while degree centrality in the insular cortex, cerebellum and precuneus (DMN) correlated with brain and gray matter volume.^18^ Correlation between LL and degree centrality of the executive and salience networks was also described.^36^ We speculate that the lack of correlation between structural imaging and *k_D_* might reflect its closer relationship with actual functional reserve that may vary even in patients with similar degree of structural damage.

### 4.7 Limitations and Future Directions

While the main strength of the study was rigorous pre-registration of all main analyses, there are also several limitations related to the fact that this study was carried out as a secondary analysis: SDMT reflects mainly cognitive processing speed and involves visual processing. Further assessments across multiple cognitive domains and sensory modalities, as well as consideration of depression as a potential confounding factor are warranted. Furthermore, our cross-sectional data cannot differentiate between state (e.g., affected by the current level of exhaustion) and trait (i.e., effect of structural damage and subsequent (mal-)adaptive plasticity) as well as premorbid patient characteristics. Normative data, assessment of test-retest reliability and longitudinal evaluation are thus needed. Future directions also include assessment of time-resolved *k_D_*, as temporal dynamics of centrality seem to provide additional insights into network dysfunction underlying CI.^20^ In case of fatigue, more fine-grained distinction into motor and cognitive fatigue would also further aid the interpretation. Finally, our data cannot be currently generalized to all individuals with MS (see our inclusion criteria), hence, a cross-validation of our results in an independent dataset is necessary before translating the results into clinical practice.

## 5. Conclusions

Although our results require further cross-validation, they suggest that obtaining a single scalar functional imaging biomarker of CI is feasible and may provide an important diagnostic tool to assess performance decline due to CI and fatigue.

## 6. Acknowledgments

None.

