## Supplementary Material for "Global Rearrangement of Degree Centrality Reflects Cognitive Impairment and Fatigue in Multiple Sclerosis"

### 1. Supplementary Methods

#### 1.1 De-noising procedure

Segmentation of the T1-weighted structural image for the denoising procedure was carried out using CAT12 Toolbox (v.12.8r1932; Christian Gaser, Jena University Hospital), yielding gray matter, white matter and cerebro-spinal fluid masks in the MNI template space. Denoising was carried out using an anatomical component-based noise correction procedure (aCompCor),[1] implemented in CONN toolbox v. 21a,[2] incorporating linear regression of noise signal extracted from the subject-specific white matter and cerebrospinal fluid masks (5 time series from principal component analysis [PCA] of each source), a regressor for each outlier volume with excessive motion (criteria: composite motion > 0.9 mm or global signal volume-to-volume change beyond 5 standard deviations [SD]), and 6 motion parameters including their 6 first-order temporal derivatives (imported from previous preprocessing steps).

#### 1.2 Voxel-wise whole-brain regions of interest (ROI)

A whole-brain voxel-wise parcellation consisted of 6-mm cubic regions of interest (ROIs) within the group-wise gray matter mask.[3] To obtain the voxel-wise parcellation with approximately 6,000 ROIs, individual gray matter segments from T1-weighted images were averaged, down-sampled to a 6-mm space with trilinear interpolation, thresholded at $p < 0.3$, and binarized. The parcellation was finally masked with a down-sampled common brain mask based on blood oxygenation level-dependent (BOLD) data, yielding 4,632 ROIs.

#### 1.3 Calculation of global degree rank order disruption index ($k_D$)

The $k_D$ was calculated using custom Matlab script (available at https://github.com/pavelhok/calculate_kd/tree/MS-project) implementing a modified approach according to Achard et al.[4] and Mansour et al.[3] To overcome the necessity for an off-site control group as in Mansour et al.,[3] we employed random sampling of a half of the control group. First, mean nodal degree (see article Section 2.5 Data pre-processing and analysis in the main manuscript body for details on degree calculation) of the control group was subtracted from the degree of the corresponding node in each participant. The difference between individual nodal degree and the control group mean was then plotted against the control group mean and $k_D$ was obtained using a linear regression (y = $k_D$ *x + b), where y = individual nodal degree – mean control group nodal degree, x = mean control group nodal degree, and b = intercept of the regression. The procedure was repeated across 100 random splittings of the control group and final $k_D$ in each patient and healthy control (HC) was calculated by averaging the $k_D$ values obtained in each iteration. For each HC participant, the final averaged $k_D$ was based on 100 splittings in which the participant was not included in the control group mean.

##### 1.4 Figure preparation

Fig. 1 was created using an open-source Python implementation of Raincould Plots available at https://github.com/pog87/PtitPrince. Fig. 2 was generated using SPSS v29.0.1.1 (IBM, Armonk, NY, USA). Plots for Fig. 3 and Fig. S4 were created in Matlab v. R2018a. Brain reconstructions and slices for Fig. 4 and Fig. S3 were prepared in Mango v. 4.1 1531 (https://rii.uthscsa.edu/mango/). Brain slices for Fig. S1 were prepared using FSLeyes v. 1.10.2 (FMRIB Centre, Oxford, UK, https://fsl.fmrib.ox.ac.uk/fsl/fslwiki/FSLeyes).

#### 2. **Supplementary Results**

##### 2.1 Study sample

Here, results including motion outliers (i.e., including 7 patients with multiple sclerosis [PwMS] and 6 healthy controls [HCs] with excessive motion levels were identified) are reported, whereas results without outliers ("final" sample) are provided in the main manuscript body. In the sample with outliers, median age in PwMS was slightly higher than in HCs (Supplementary Table 5).

##### 2.2 Group differences and group differentiation (hypotheses 1 and 2)

PwMS showed significantly lower degree rank order disruption index ($k_D$) compared to HCs (PwMS: median = -0.316, inter-quartile range [IQR] = 0.498; HCs: median = -0.082, IQR = 0.541; $p = 0.001$, Mann-Whitney U test).

For hypothesis 2a, the receiver operating characteristic (ROC) analysis for differentiation between PwMS and HCs yielded significant above-chance area under curve (AUC) for $k_D$ (AUC = 0.667, $p$ = 0.001, two-tailed asymptotic significance for null hypothesis AUC = 0.5), the left lateral parietal portion of the DMN (DMN-LLP; AUC = 0.677, $p < 0.001$), left hippocampus (AUC = 0.608, $p = 0.032$) and the ACC (AUC = 0.606; $p = 0.036$), see Supplementary Table 6. In pair-wise comparisons, AUC for $k_D$ was significantly higher than AUC for 11 ROIs and did not significantly differ from the remaining ROIs (Supplementary Table 6).

For hypothesis 2b, we observed no significant improvement in a multiple logistic regression model differentiating between PwMS and HCs) after adding $k_D$ as an additional regressor on top of gray matter volume (GMV), fractional anisotropy (FA), log(lesion load [LL]) ($\chi^2$ step = 0.579, $p = 0.447$).

##### 2.3 Correlation with cognitive processing speed (hypotheses 3 and 4)

We detected no significant correlation between $k_D$ and SDMT (Spearman's $\varrho = 0.20$, $p = 0.111$, $n = 62$). In case of regional degree centrality (hypothesis 4a), no significant correlation was observed after correction for multiple comparisons, see Supplementary Table 7. For hypothesis 4b, an ordinal regression model including GMV, FA, log(LL), age, gender, and years since diagnoses as Symbol Digit

Modalities Test (SDMT) score as regressors was not significantly improved after adding $k_D$ ($\chi^2$ step = 3.63, $p$ = 0.057, likelihood ratio test, see Supplementary Table 8).

#### 2.4 Correlation with global disability, fatigue, and motor performance (exploratory hypotheses 5 and 6)

We detected a significant correlation between $k_D$ and Fatigue Scale for Motor and Cognitive Functions score (FSMC; Spearman's $\varrho$ = –0.27, $p$ = 0.030, $n$ = 63), but not for Expanded Disability Status Scale score (EDSS; Spearman's $\varrho$ = –0.08, $p$ = 0.546, $n$ = 63) or Timed Up and Go Test (TUG; Spearman's $\varrho$ = –0.16, $p$ = 0.233, $n$ = 58). For hypothesis 6, $k_D$ significantly improved an ordinal regression model including GMV, FA, log(LL), age, gender, and years since diagnoses as regressors of fatigue (FSMC), but not for EDSS or TUG (Supplementary Table 8).

#### 2.5 Relationship between $k_D$ and structural imaging biomarkers (exploratory hypotheses 7 and 8)

We observed a significant correlation (hypothesis 7) between $k_D$ and LL (Spearman's $\varrho$ = –0.27, $p$ = 0.033, $n$ = 63), but no significant correlation with GMV (Spearman's $\varrho$ = 0.12, $p$ = 0.354, $n$ = 63) or global FA (Spearman's $\varrho$ = 0.04, $p$ = 0.731, $n$ = 63). All structural imaging parameters significantly differed between PwMS and HCs (hypothesis 8), see Supplementary Table 9.

#### 3. Supplementary Tables

**Supplementary Table 1. List of regions of interests (ROI)**

| Abbreviation | Description | Side | MNI coordinates[a] (x, y, z) [mm] | Size [voxels][b] | Source[c] |
|---|---|---|---|---|---|
| **DMN-MPFC** | default mode network, medial prefrontal cortex | | 1, 52, -3 | 34 | CONN network atlas[2] binary labels |
| **DMN-LP** | default mode network, lateral parietal part | L | -40, -76, 32 | 32 | |
| | | R | 47, -66, 29 | 42 | |
| **DMN-PCC** | default mode network, posterior cingulate cortex | | 1, -61, 37 | 161 | |
| **Put** | putamen | L | -25, 0, 1 | 32 | HOSA[5–8] 25% maximum probability labels |
| | | R | 26, 2, 1 | 29 | |
| **Cau** | caudate nucleus | L | -13, 10, 10 | 15 | |
| | | R | 15, 11, 11 | 20 | |
| **GP**[d] | globus pallidus | L | -24, -6, -6 | 1 | |
| | | R | 18, 6, 0 | 1 | |
| **Tha** | thalamus | L | -8, -20, 7 | 37 | |
| | | R | 11, -20, 8 | 38 | |
| **Hip** | hippocampus | L | -27, -21, -15 | 21 | |
| | | R | 28, -21, -14 | 24 | |
| **Crbl** | cerebellum | | 2, -61, -31 | 461 | MNI structural atlas[9,10] 25% maximum probability labels |
| **SPL** | superior parietal lobule | L | -18, -63, 57 | 18 | Spherical ROI (d = 18 mm) centered according to Grothe et al.[11] |
| | | R | 21, -66, 51 | 17 | |
| **DLPFC** | dorsolateral prefrontal cortex | L | -24, -3, 51 | 9 | |
| | | R | 33, 0, 60 | 13 | |
| **ACC** | anterior cingulate cortex | | 9, 15, 39 | 9 | |

Notes: [a)]Atlas ROIs: coordinates are centers of mass of final ROIs, spherical ROIs: coordinates are centers of original spheres; [b)]voxel size 6×6×6 mm; [c)]All ROIs were additionally masked with common gray matter and functional brain mask; [d)]region excluded from analyses due to small size after resampling.
Abbreviations: HOSA – Harvard-Oxford subcortical atlas, L – left; MNI – Montreal Neurological Institute; R – right; ROI – region of interest.

**Supplementary Table 2. Summary of outcome measures, regressors and statistical tests**

| Number | Hypothesis | Outcome measures | Regressors | Confounders | Statistical Test |
|---|---|---|---|---|---|
| **1** | Group differences in $k_D$ | $k_D$ | Presence of MS | none | Mann-Whitney U test |
| **2** | Differentiation between PwMS and HCs | Presence of MS | $k_D$, regional degree from 18 ROIs | none | ROC analysis with AUC pairwise comparisons |
| | | | $k_D$, GMV, FA, log(LL) | none | multiple logistic regression with likelihood ratio test |
| **3** | Correlation with cognitive processing speed | SDMT | $k_D$ | none | Spearman's rank correlation coefficient |
| **4** | Regression of cognitive processing speed | SDMT | regional degree from 18 ROIs | none | Spearman's rank correlation coefficient |
| | | | $k_D$, GMV, FA, log(LL) | age, sex, years since diagnosis | ordinal regression with likelihood ratio test |
| | | | **Exploratory hypotheses** | | |
| **5** | Correlation with global disability, fatigue, and motor performance | FSMC, EDSS, TUG | $k_D$ | none | Spearman's rank correlation coefficient |
| **6** | Regression of global disability, fatigue, and motor performance | FSMC, EDSS, TUG | $k_D$, GMV, FA, log(LL) | age, sex, years since diagnosis | ordinal regression with likelihood ratio test |
| **7** | Relationship between $k_D$ and structural imaging biomarkers | $k_D$ | GMV, FA, log(LL) | none | Spearman's rank correlation coefficient |
| **8** | Group differences in structural imaging biomarkers | GMV, FA, log(LL) | Presence of MS | none | Mann-Whitney U test |

Abbreviations: EDSS – Expanded Disability Status Scale; FA – fractional anisotropy; FSMC – Fatigue Scale for Motor and Cognitive Functions; GMV – gray matter volume; HCs – healthy controls; $k_D$ – degree rank order disruption index; log(LL) – log(lesion load); MS – multiple sclerosis; PwMS – patients with MS; ROIs – regions of interest; SDMT – Symbol Digit Modalities Test; TUG – Timed Up and Go Test.

**Supplementary Table 3. Receiver operating characteristic (ROC) analysis for group membership – no outliers**

| ROI | | AUC | $p^a$ | AUC difference[b] | $p^c$ |
|---|---|---|---|---|---|
| **DMN-MPFC** | | 0.570 | 0.197 | 0.072 | 0.116 |
| **DMN-LP** | **L** | **0.671** | **0.001** | -0.029 | 0.585 |
| | **R** | 0.572 | 0.188 | 0.070 | 0.222 |
| **DMN-PCC** | | 0.510 | 0.860 | 0.132 | 0.128 |
| **Put** | **L** | 0.540 | 0.467 | **0.102** | **0.004** |
| | **R** | 0.547 | 0.384 | **0.095** | **0.008** |
| **Cau** | **L** | 0.573 | 0.182 | **0.069** | **0.034** |
| | **R** | 0.584 | 0.118 | 0.058 | 0.054 |
| **Tha** | **L** | 0.580 | 0.140 | 0.062 | 0.171 |
| | **R** | 0.571 | 0.191 | 0.071 | 0.139 |
| **Hip** | **L** | 0.560 | 0.267 | 0.082 | 0.075 |
| | **R** | 0.534 | 0.535 | **0.108** | **0.030** |
| **Crbl** | | 0.526 | 0.628 | **0.115** | **<0.001** |
| **SPL** | **L** | 0.509 | 0.864 | 0.133 | 0.123 |
| | **R** | 0.502 | 0.966 | **0.140** | **0.030** |
| **DLPFC** | **L** | 0.506 | 0.917 | 0.136 | 0.059 |
| | **R** | 0.555 | 0.311 | 0.087 | 0.314 |
| **ACC** | | **0.619** | **0.026** | 0.023 | 0.619 |

**Notes:** [a)]Asymptotic one-tailed uncorrected $p$ for null hypothesis: true area = 0.5, significant values at $p < 0.05$ marked in **bold**; [b)]$AUC_{kD}$ – AUC; [c)]Asymptotic two-tailed uncorrected $p$ for null hypothesis: true area difference = 0, significant values at $p < 0.05$ marked in **bold**.

**Abbreviations:** ACC – anterior cingulate cortex; AUC – area under curve; Cau – caudate nucleus; Crbl – cerebellum; DLPFC – dorsolateral prefrontal cortex; DMN – default mode network: -LP – lateral parietal part, -MPFC – medial prefrontal cortex, -PCC – posterior cingulate cortex; Hip – hippocampus; L – left; Put – putamen; SPL – superior parietal lobule; Tha – thalamus; R – right; ROI – region of interest.

**Supplementary Table 4. Group differences in structural imaging parameters – no outliers**

| | PwMS $n = 56$ | HC $n = 58$ | |
|---|---|---|---|
| | **Median ±IQR** | **Median ±IQR** | $p^a$ |
| **Lesion load [mm³]** | 2682.7 ±4834.4 | 91.0 ±114.2 | **<0.001** |
| **log(Lesion load) [log(mm³)]** | 3.43 ±0.81 | 1.96 ±0.52 | |
| **Gray matter volume [cm³]** | 1410.3 ±236.4 | 1591.6 ±195.0 | **<0.001** |
| **Global FA** | 0.588 ±0.033 | 0.612 ±0.029 | **<0.001** |

**Notes:** [a)]Mann-Whitney U Test.

**Abbreviations:** FA – fractional anisotropy; HC – healthy controls; IQR – interquartile range; PwMS – patients with MS.

**Supplementary Table 5 Demographic and clinical data – sample with and without outliers**

| | | Enrolled subjects | Included subjects | |
|---|---|---|---|---|
| | | | With outliers | Final sample |
| **HC** | | | | |
| **Number** | | 65 | 64 | 58 |
| **Gender [women/men]** | | 32/33 | 31/33 | 27/31 |
| **Median age ±IQR [yrs]** | | 40.9 ±17 | 40.8 ±17 | 40.5 ±17 |
| **PwMS** | | | | |
| **Number** | | 65 | 63 | 56 |
| **Gender [women/men]** | | 39/26 | 38/25 | 35/21 |
| $p^a$ | | 0.291 | 0.214 | 0.095 |
| **Median age ±IQR [yrs]** | | 45.3 ±17 | 45.4 ±17 | 45.1 ±17 |
| $p^b$ | | **0.048** | **0.039** | 0.090 |
| **Diagnosis [n, %]** | **RRMS** | 38, 58.5% | 38, 60.3% | 35, 62.5% |
| | **SPMS** | 20, 30.8% | 18, 28.6% | 15, 26.8% |
| | **PPMS** | 6, 9.2% | 6, 9.5% | 5, 8.9% |
| | **no data** | 1, 1.5% | 1, 1.6% | 1, 1.8% |
| **Time since diagnosis ±SD [yrs]** | | 12.8 ±6.8 | 12.7 ±6.9 | 12.6 ±6.2 |
| **EDSS ±IQR** | | 4.5 ±2.0 | 4.5 ±2.0 | 4.5 ±3.0 |
| **SDMT ±IQR** | | 45 ±31 | 45 ±31 | 45 ±29 |
| **FSMC ±IQR** | | 57 ±23 | 57 ±23 | 57 ±23 |
| **TUG ±IQR [s]** | | 11.2 ±11 | 10.8 ±9 | 10.3 ±9 |

**Notes:** [a)] Fisher's exact test between PwMS and HCs; [b)] Mann-Whitney U test between PwMS and HCs.

**Abbreviations:** EDSS – Expanded Disability Status Scale; FSMC – Fatigue Scale for Motor and Cognitive Functions; HCs – healthy controls; IQR – interquartile range; MS – multiple sclerosis; n – number; N/A – not applicable; PPMS – primary progressive MS; PwMS – patients with MS; RRMS – relapsing-remitting MS; SD – standard deviation; SDMT – Symbol Digit Modalities Test; SPMS – secondary progressive MS; TUG – Timed Up and Go Test; yrs – years.

**Supplementary Table 6. Receiver operating characteristic (ROC) analysis for group membership – with outliers**

| ROI | | AUC | $p^a$ | AUC difference[b] | $p^c$ |
|---|---|---|---|---|---|
| **DMN-MPFC** | | 0.585 | 0.094 | 0.082 | 0.060 |
| **DMN-LP** | **L** | 0.677 | **<0.001** | -0.010 | 0.835 |
| | **R** | 0.586 | 0.094 | 0.081 | 0.132 |
| **DMN-PCC** | | 0.526 | 0.621 | 0.141 | 0.079 |
| **Put** | **L** | 0.558 | 0.257 | 0.109 | **0.001** |
| | **R** | 0.573 | 0.156 | 0.094 | **0.004** |
| **Cau** | **L** | 0.583 | 0.103 | 0.083 | **0.006** |
| | **R** | 0.587 | 0.089 | 0.080 | **0.005** |
| **Tha** | **L** | 0.581 | 0.115 | 0.086 | **0.041** |
| | **R** | 0.575 | 0.146 | 0.092 | **0.042** |
| **Hip** | **L** | 0.608 | **0.032** | 0.059 | 0.152 |
| | **R** | 0.576 | 0.139 | 0.091 | **0.045** |
| **Crbl** | | 0.531 | 0.547 | 0.136 | **<0.001** |
| **SPL** | **L** | 0.512 | 0.814 | 0.155 | **0.008** |
| | **R** | 0.522 | 0.675 | 0.145 | **0.012** |
| **DLPFC** | **L** | 0.504 | 0.937 | 0.163 | **0.018** |
| | **R** | 0.572 | 0.162 | 0.095 | 0.230 |
| **ACC** | | 0.606 | **0.036** | 0.061 | 0.168 |

**Notes:** [a)]Asymptotic one-tailed uncorrected *p* for null hypothesis: true area = 0.5, significant values at $p < 0.05$ marked in **bold**; [b)]$AUC_{kD}$ – AUC; [c)]Asymptotic two-tailed uncorrected *p* for null hypothesis: true area difference = 0, significant values at $p < 0.05$ marked in **bold**.

**Abbreviations:** ACC – anterior cingulate cortex; AUC – area under curve; Cau – caudate nucleus; Crbl – cerebellum; DLPFC – dorsolateral prefrontal cortex; DMN – default mode network: -LP – lateral parietal part, -MPFC – medial prefrontal cortex, -PCC – posterior cingulate cortex; Hip – hippocampus; L – left; Put – putamen; SPL – superior parietal lobule; Tha – thalamus; R – right; ROI – region of interest.

**Supplementary Table 7. Correlation between regional degree and clinical scores – with outliers**

| | | SDMT $n$ = 62 | | FSMC $n$ = 63 | |
|---|---|---|---|---|---|
| | | $\varrho^a$ | $p^a$ | $\varrho^a$ | $p^a$ |
| **DMN-MPFC** | | -0.354 | *0.005* | 0.200 | 0.116 |
| **DMN-LP** | **L** | 0.062 | 0.635 | 0.098 | 0.443 |
| | **R** | -0.129 | 0.318 | 0.251 | *0.047* |
| **DMN-PCC** | | -0.169 | 0.189 | 0.152 | 0.233 |
| **Put** | **L** | -0.200 | 0.119 | 0.326 | *0.009* |
| | **R** | -0.114 | 0.376 | 0.254 | *0.044* |
| **Cau** | **L** | -0.184 | 0.153 | 0.341 | *0.006* |
| | **R** | -0.199 | 0.121 | 0.323 | *0.010* |
| **Tha** | **L** | -0.149 | 0.247 | 0.240 | 0.058 |
| | **R** | -0.191 | 0.137 | 0.229 | 0.071 |
| **Hip** | **L** | -0.170 | 0.188 | 0.172 | 0.178 |
| | **R** | -0.306 | *0.016* | 0.236 | 0.063 |
| **Crbl** | | -0.243 | 0.057 | 0.357 | *0.004* |
| **SPL** | **L** | -0.112 | 0.384 | -0.131 | 0.304 |
| | **R** | -0.170 | 0.186 | -0.041 | 0.748 |
| **DLPFC** | **L** | -0.143 | 0.266 | -0.157 | 0.220 |
| | **R** | -0.111 | 0.388 | 0.068 | 0.595 |
| **ACC** | | -0.104 | 0.421 | 0.006 | 0.960 |

**Notes:** [a)]Spearman's rank correlation coefficient $\varrho$, significant correlations at Bonferroni-Holm-corrected alpha = 0.0028 are marked in **bold** type, significant correlations at uncorrected alpha = 0.05 are marked in *italics*.

**Abbreviations:** ACC – anterior cingulate cortex; Cau – caudate nucleus; Crbl – cerebellum; DLPFC – dorsolateral prefrontal cortex; DMN – default mode network: -LP – lateral parietal part, -MPFC – medial prefrontal cortex, -PCC – posterior cingulate cortex; FSMC – Fatigue Scale for Motor and Cognitive Functions; Hip – hippocampus; L – left; n – number; Put – putamen; SPL – superior parietal lobule; Tha – thalamus; R – right; ROI – region of interest; SDMT – Symbol Digit Modalities Test.

**Supplementary Table 8. Ordinal regression of clinical scores – with outliers**

| Regressand | SDMT | | EDSS | | FSMC | | TUG | |
|---|---|---|---|---|---|---|---|---|
| **Model** | **No $k_D$** | **With $k_D$** | **No $k_D$** | **With $k_D$** | **No $k_D$** | **With $k_D$** | **No $k_D$** | **With $k_D$** |
| **Pseudo $R^2$ (Cox&Snell)** | 0.230 | 0.274 | 0.320 | 0.321 | 0.125 | 0.212 | 0.296 | 0.301 |
| **-2 Log Likelihood** | 433.806 | 430.179 | 282.218 | 282.123 | 439.047 | 432.515 | 442.300 | 441.883 |
| $\chi^2$ | 16.227 | 19.855 | 24.333 | 24.427 | 8.441 | 14.973 | 20.394 | 20.810 |
| **df** | 6 | 7 | 6 | 7 | 6 | 7 | 6 | 7 |
| **Model Sig.** | **0.013** | **0.006** | **<0.001** | **0.001** | 0.208 | **0.036** | **0.002** | **0.004** |
| **$k_D$ Wald** | N/A | 3.307 | N/A | 0.111 | N/A | 6.637 | N/A | 0.454 |
| **$k_D$ Sig.** | N/A | 0.069 | N/A | 0.739 | N/A | **0.010** | N/A | 0.500 |
| **$\chi^2$ step** | 3.63 | | 0.09 | | 6.53 | | 0.42 | |
| **df** | 1 | | 1 | | 1 | | 1 | |
| **$p^a$** | 0.057 | | 0.759 | | **0.011** | | 0.519 | |

**Notes:** [a)]One-tailed likelihood ratio test.

**Abbreviations:** df – degrees of freedom; EDSS – Expanded Disability Status Scale; FSMC – Fatigue Scale for Motor and Cognitive Functions; $k_D$ – degree rank order disruption index; n – number; N/A – not applicable; SDMT – Symbol Digit Modalities Test; TUG – Timed Up and Go Test.

**Supplementary Table 9. Group differences in structural imaging parameters – with outliers**

| | PwMS $n = 63$ | HC $n = 64$ | |
|---|---|---|---|
| | **Median ±IQR** | **Median ±IQR** | $p^a$ |
| **Lesion load [mm³]** | 2765.2 ±5115.6 | 93.7 ±134.9 | **<0.001** |
| **log(Lesion load) [log(mm³)]** | 3.44 ±0.83 | 1.97 ±0.58 | |
| **Gray matter volume [cm³]** | 1399.8 ±250.0 | 1561.3 ±193.5 | **<0.001** |
| **Global FA** | 0.588 ±0.039 | 0.610 ±0.028 | **<0.001** |

**Notes:** [a)]Mann-Whitney U Test.

**Abbreviations:** FA – fractional anisotropy; HC – healthy controls; IQR – interquartile range; PwMS – patients with MS.

#### 4. Supplementary Figures

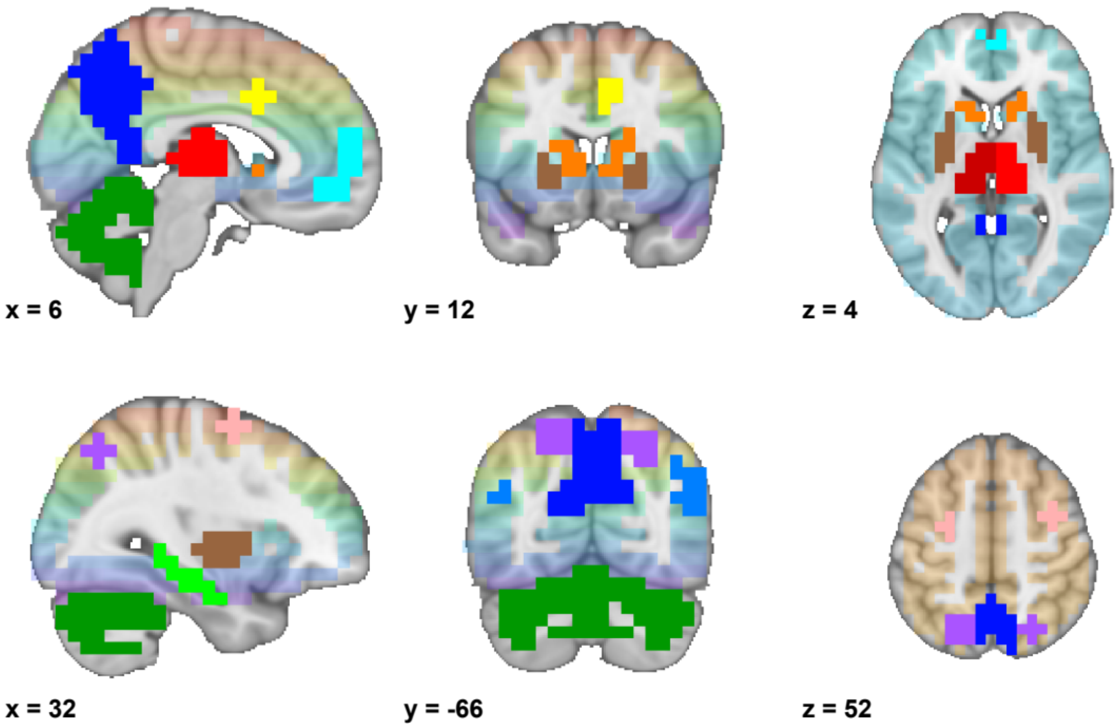


**Supplementary Fig. S1. Regions of interest (ROIs).** Color overlays representing ROIs on top of orthogonal slices of the MNI152 standard brain template. Color coding: color spectrum (transparent background) – included 6-mm voxels; cyan - default mode network, medial prefrontal cortex; light blue - default mode network, lateral parietal cortex; dark blue – default mode network, posterior cingulate cortex; brown – putamen; orange – caudate nucleus; red (light & dark) – thalamus; light green – hippocampus; dark green – cerebellum; purple – superior parietal lobule; pink – dorsolateral prefrontal cortex; yellow – anterior cingulate cortex.

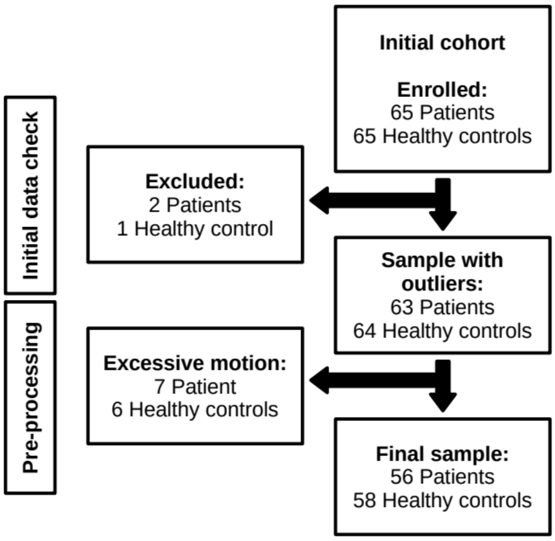


**Supplementary Fig. S2. Inclusion/exclusion diagram.** Diagram illustrates exclusion rates at each step of the data analysis.

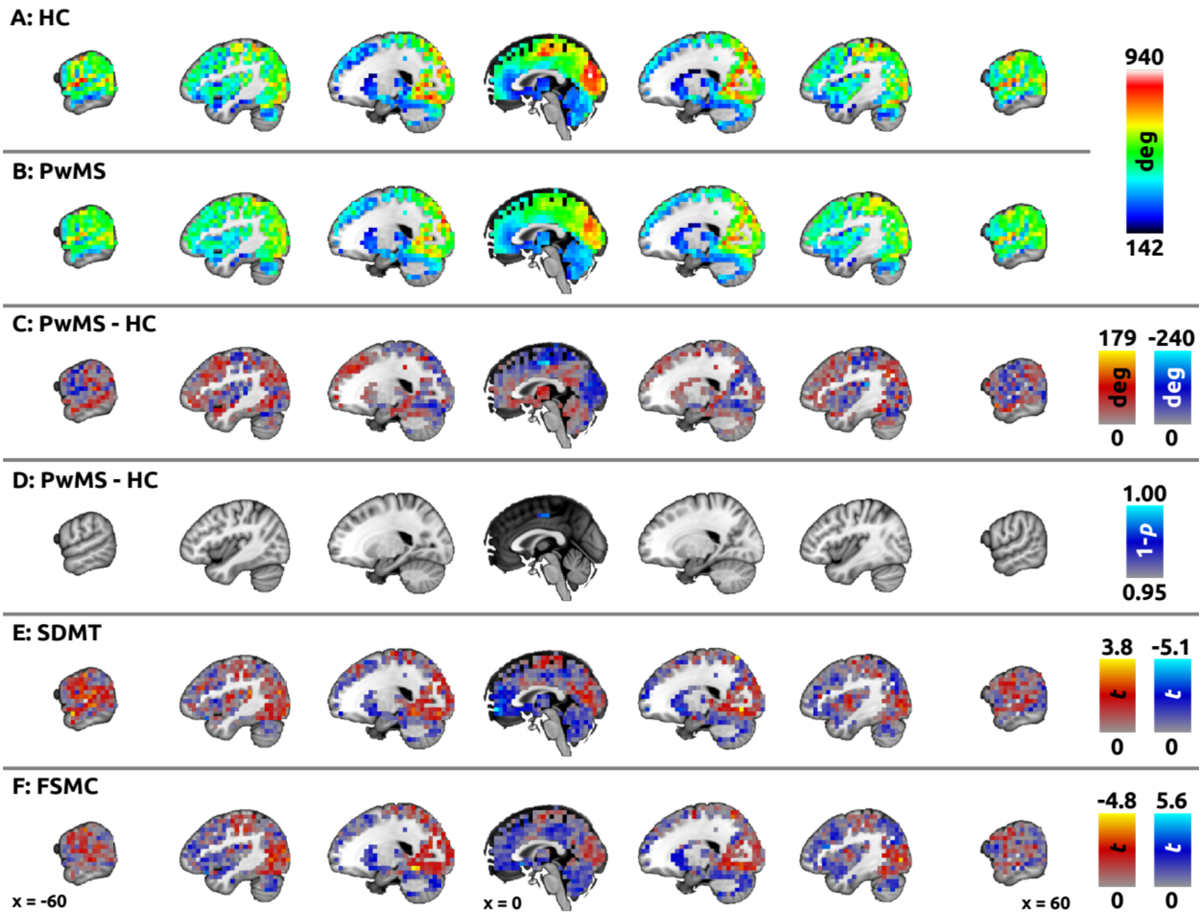


**Supplementary Fig. S3. Raw degree centrality, group degree differences, and unthresholded data.** Color overlays on top of 1-mm MNI152 standard brain sagittal slices illustrate the underlying data for main analyses. In **panel A**, mean raw degree in healthy control (HC) group is shown (no outliers, $n = 58$). **Panel B** shows mean raw degree in patients with multiple sclerosis (PwMS, $n = 56$), using the same color scaling (actual range for PwMS = 172-843). **Panel C** shows mean difference individual degree in PwMS – mean normal degree in HC ($n = 56$), red overlay indicates higher degree in PwMS, blue overlay indicates higher degree in HC. In **panel D**, statistically significant group differences in raw degree are shown (thresholded using non-parametric threshold-free cluster enhancement with 10,000 permutations, family-wise error-corrected $p = 0.05$), with blue overlay indicating higher degree in HC in supplementary motor area and adjacent paracentral lobule. **Panels E-F** show unthresholded t-maps illustrating spatial distribution of linear regression of the degree centrality for (**E**) cognitive processing speed (Symbol Digit Modalities Test, SDMT) and (**F**) fatigue (Fatigue Scale for Motor and Cognitive Functions, FSMC). Here, color-coding was inverted for FSMC (positive correlation in blue, negative correlation in red) to match color coding for SDMT (in general, impairment is associated with lower SDMT, but higher FSMC).

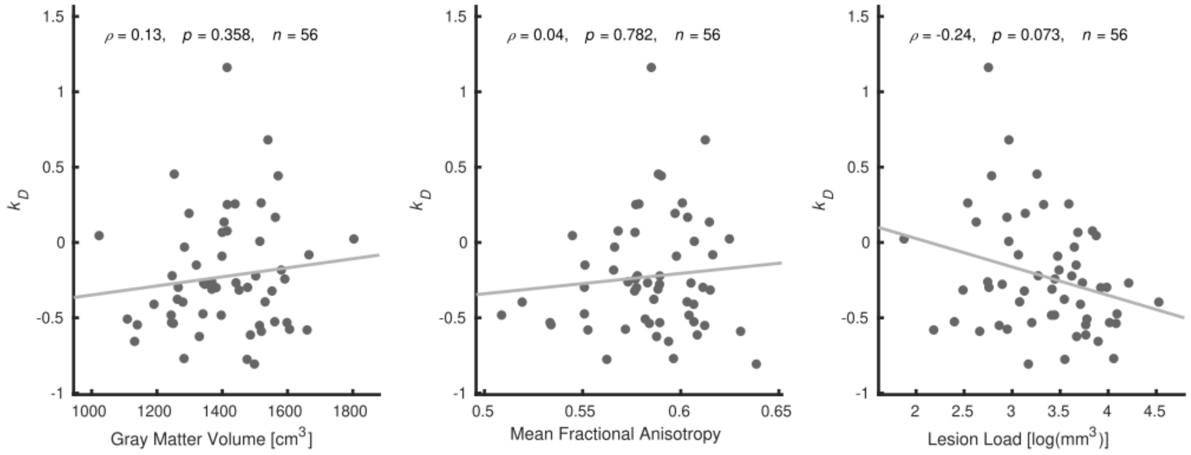


**Supplementary Fig. S4. Correlation between *$k_D$* and structural imaging.** Scatter plots illustrating relationship between the degree rank order disruption index ($k_D$) and global gray matter volume, global white matter fractional anisotropy, and lesion load (after log transform). Spearman's rank correlation coefficient ($\varrho$), two-tailed uncorrected significance, and number of valid observations are provided.
